# Pleuroparenchymal fibroelastosis in monogenic *DGUOK*-associated mitochondriopathy

**DOI:** 10.64898/2026.04.08.26349275

**Authors:** Sandra von Hardenberg, Pia Maier, Leonard Christian, Anibh Martin Das, Lavinia Neubert, Jannik Ruwisch, Katrin Peters, Dirk Schramm, Matthias Griese, Britta Skawran, Marlies Eilers, Danny Jonigk, Norman Junge, Aiden Haghikia, Tobias Hegelmaier, Winfried Hofmann, Benjamin Seeliger, Diane Miriam Renz, Amelie Stalke, Lara Hartmayer, Alexander Duscha, Martin Schulze, Nataliya DiDonato, Holger Prokisch, Bernd Auber, Lars Knudsen, Jonas C Schupp, Nicolaus Schwerk

## Abstract

**Background:** Pleuroparenchymal fibroelastosis (PPFE) is a rare, fibrotic lung disease with poor prognosis, usually affecting adults which most commonly occurs idiopathically. Biallelic pathogenic variants in *DGUOK* cause mitochondrial DNA (mtDNA) depletion syndrome, predominantly affecting infants with severe hepatic and neurological symptoms. Detailed description of pulmonary manifestations with late-onset presentation have not been reported.

**Methods:** We describe nine patients with PPFE and *DGUOK*-associated mitochondriopathy. Clinical, radiological, histopathological, and genetic data were systematically collected from all patients. Functional studies, single nucleus RNA sequencing (snRNAseq), immunofluorescence staining, transmission electron microscopy and respiratory chain enzyme activity assays were conducted on patient-derived fibroblasts, muscle or lung tissues. mtDNA content quantification was performed on whole genome sequencing (WGS) data.

**Results:** All patients (ages 5–36) presented with progressive dyspnea, weight loss and some with spontaneous pneumothoraces. Chest computed tomography and lung biopsies showed features of PPFE. Biallelic pathogenic *DGUOK* variants were identified in all patients, seven of them carry an unreported intronic variant leading to mtDNA depletion. snRNAseq of lung tissue from four pediatric patients identified Aberrant Basaloid cells and intermediate cells as their precursor localized at the fibrotic edge. Mitochondrial alterations were identified by electron microscopy.

**Conclusion:** PPFE in children and young adults is associated with *DGUOK*-related mitochondriopathy. For the first time, we demonstrate Aberrant Basaloid cells in pediatric fibrotic lung tissue. Since pulmonary involvement may be underrecognized or misinterpreted and the clinical presentation may not always be typical of a mitochondriopathy, we recommend genetic testing in all patients with PPFE of unknown origin.

## Introduction

Pleuroparenchymal fibroelastosis (PPFE) is a rare fibrotic lung disease histologically characterized by subpleural alveolar fibroelastosis, usually with upper lobe predominance^1^. Typical clinical findings are progressive dyspnea, chronic nonproductive cough, weight loss, platythorax, deep sternal notch and a restrictive ventilatory defect^2,3^. After the onset of clinical symptoms, the disease often progresses rapidly, which is reflected in a poor prognosis with 5-year survival rates ranging from 20-60%^4,5^

PPFE is schematically categorized as idiopathic (iPPFE) or secondary (sPPFE) due to different conditions^2^. iPPFE is categorized as a specific disease entity among other idiopathic interstitial pneumonias (IIP) in the American Thoracic Society/European Respiratory Society (ATS/ERS) statement on the classification of IIPs. While this disorder has been increasingly described in adults, there are only very few children reported, mostly after chemotherapy or following bone marrow transplantation^3,6,7^. Since the first description in 2004, growing evidence from familial cases of iPPFE and the detection of pathogenic variants in telomerase genes in some patients suggests a hereditary contribution^1,6,8–13^. However, to date, no specific monogenetic cause has been assigned to iPPFE and there is currently no evidence as to the etiology and pathophysiology of this disease.

There is growing evidence that acquired mitochondrial dysfunction contributes to the pathogenesis of idiopathic pulmonary fibrosis^14–17^. However, no primary causal link between mitochondrial dysfunction and the development of fibrotic lung disease has yet been identified in humans. Deoxyguanosine kinase (DGUOK) is a key enzyme of the mitochondrial deoxyribonucleoside triphosphate (dNTP) salvage pathway, required for mitochondrial DNA (mtDNA) synthesis^18^. Reduced or absent DGUOK activity disrupts the mitochondrial dNTP pool, leading to mtDNA depletion and/or multiple mtDNA deletions^19^. DGUOK deficiency causes an autosomal recessive mtDNA depletion syndrome, classically presenting in newborns and infants with severe hepatic and neurological involvement^20,21^. In addition, very rare cases of isolated hepatic disease in children and late-onset myopathy have been described^21,22^. In a recently published review of 173 patients with DGUOK deficiency, respiratory involvement was reported in 13% of cases; however, the underlying causes were not described^23^. Additionally, an unspecified interstitial lung disease was reported in one patient in a case series of 24 individuals^24^, and a pneumothorax in a separate case report^25^. Furthermore, an increasing number of patients with pulmonary hypertension have been reported^21,26^. Here, we describe five pediatric and four adult patients with PPFE and a distinct phenotype compared to ‘classical’ features of DGUOK deficiency. Our findings support the relevance of mitochondrial dysfunction in the pathophysiology of PPFE and highlight the importance of a holistic diagnostic evaluation including genetic workup in patients presenting with diffuse lung diseases, especially when these are considered idiopathic.

## Material and Methods

### Study information

Written informed consent was obtained from the patients or their legal guardians at the respective institutions. The study was approved by the Ethics Committee (Ethical vote 2923-2015, 1969-2013 and 12241-BO-K-2026) at Hannover Medical School.

### Genetic and Functional Analyses

Details of genetic, transcriptomic and cellular analyses are provided in the Supplementary Appendix.

## Results

### Clinical phenotype

A total of nine patients from four families were included (consanguinities are shown in Figure 1, and Table S3, Supplementary Appendix). P1, P2, and P3 originate from one family (F1), with P1 and P2 being siblings and P3 a distant cousin. P4 and P5 come from different families (F2 and F3), while P6–P9 are siblings (F4) (Figure 1A–D). Age at onset of respiratory symptoms varied between 1 and 28 years. At time of first presentation, leading features were dyspnea, chronic cough, a deepened suprasternal notch (Figure 1), platythorax and low body weight. All patients had a restrictive ventilatory defect. Four patients presented with a spontaneous pneumothorax. Chest-CT’s showed typical features of PPFE in all subjects (Figure 1, Table 1). Lung biopsies, available from four pediatric patients (P1-P4) revealed alveolar fibroelastosis (AFE), the histological pattern underlying PPFE (Figure 1I, J). In those, treatment with methylprednisolone-pulses, prednisolone and nintedanib was attempted, without apparent benefit and all discontinued nintedanib due to side effects (nausea, vomiting, and further weight loss). Two patients underwent liver transplantation within the first three years of life due to acute liver failure. In contrast to the other cases, these two showed psychomotor developmental delay over time. All other patients did not have clinical signs of liver disease or neurodevelopmental delay, even though liver enzymes were slightly elevated in five non-liver transplanted patients. Extrapulmonary organ manifestations were fatigue (n=4), muscle weakness (n=6), peripheral polyneuropathy with predominantly axonal damage (n=5), proteinuria (n=4). P1 underwent lung transplantation (LTx) seven months after diagnosis. P3 and P4 were also evaluated for LTx due to rapid respiratory deterioration but were considered unsuitable for Ltx because of severe cachexia and rapidly progressive polyneuropathy with complete immobilization. P3 died 1.5 years after onset of respiratory symptoms and six months after PPFE-diagnosis due to terminal respiratory failure. P5 was not evaluated for LTx and died before the age of six due to cardio-respiratory failure. The remaining six patients are still alive at last follow-up. Representative radiological, histological and clinical images are shown in Figure 1. Table 1 and Table S3, Supplementary Appendix, provide a detailed overview of the symptoms and findings.

**Figure 1.**
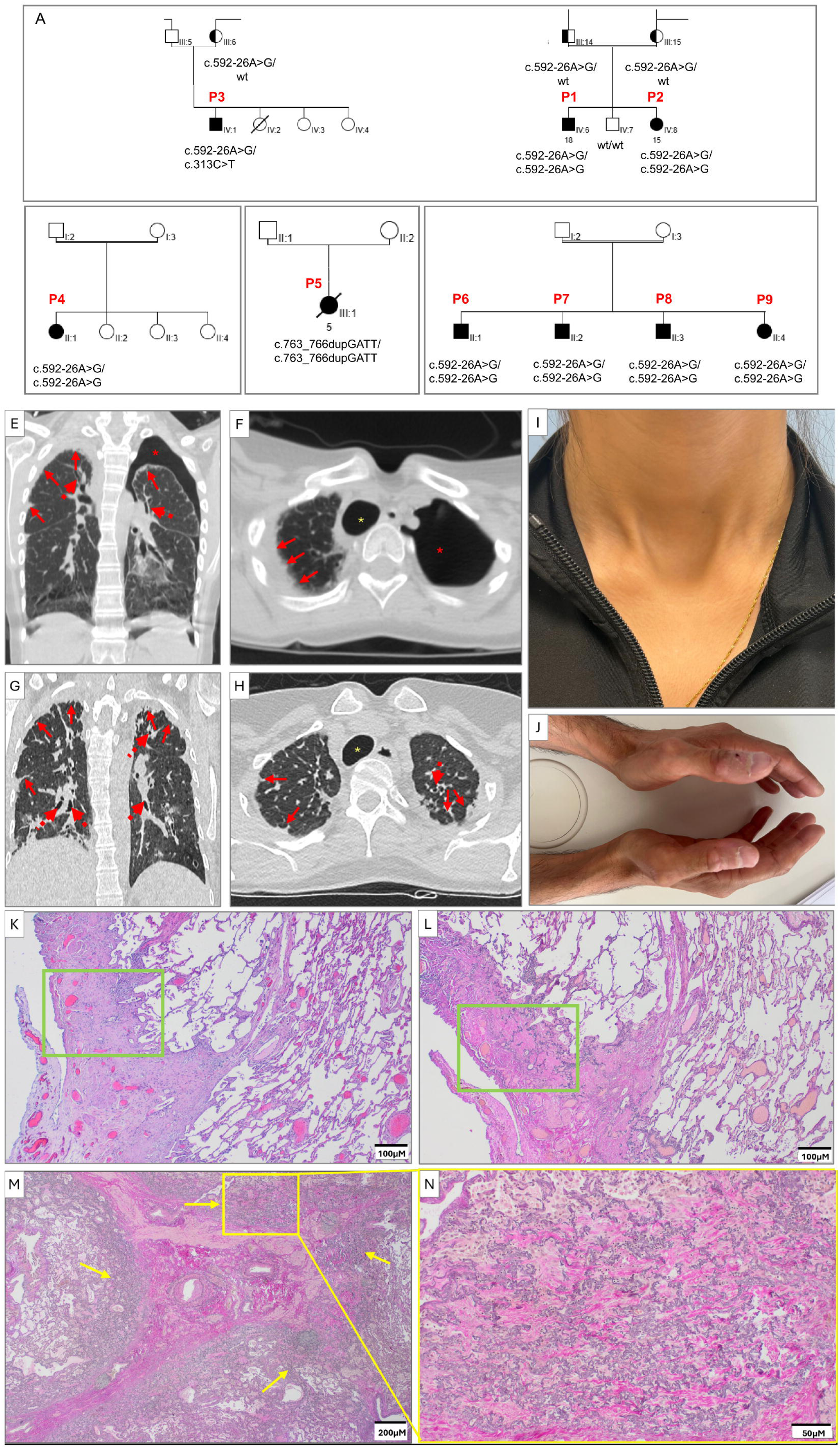
Genetic and clinical features of patients with DGUOK-associated mitochondriopathy. Panel A-D shows the pedigrees of four unrelated families. Double lines denote consanguinity, filled symbols indicate affected individuals with biallelic *DGUOK* variants and half-filled symbols indicate heterozygous carriers. Genotypes are shown below the symbols. Squares and circles represent males and females, respectively. A slash denotes deceased individuals. Roman numerals indicate generations, whereas Arabic numerals denote individuals within each generation. Representative chest CT images of patients are presented in Panel E-H with P4 (E, F) and P6 (G, H), showing predominantly apical, subpleural and septal thickening (red arrows), traction bronchiectasis (red dashed arrows), pneumothorax (red asterisk) and tracheal deviation (yellow asterisk). Panel I shows a deepened suprasternal notch, characteristic for fibrotic contraction of the upper lung lobes in P2. The hands of the patients show no signs of digital clubbing (P6) but severe atrophy of the small hand muscles (Panel J). Panels K–N illustrate the characteristics and rapid progression of PPFE in patient P1. At diagnosis, Panels K and L show a video-assisted thoracoscopic surgery (VATS) biopsy with Hematoxylin and Eosin (HE) and Elastica van Gieson (EvG) staining, respectively, revealing a sharply demarcated subpleural area with an alveolar fibroelastosis (AFE) pattern, highlighted by green boxes. Panel L illustrates the characteristic AFE features, including prominent black-stained elastic fibers in the areas of the former alveolar walls and the former alveolar lumina filled with pink-stained collagen. Panels M and N show HE and EvG staining of the explanted lung obtained 8 months later, demonstrating advanced fibrotic remodeling, highlighted by yellow arrows.

**Table 1.** Demographic and Clinical Characteristics of Nine Patients with DGUOK Deficiency.

| Characteristics | Patients (n=9) |
| --- | --- |
| **Age** |  |
| <10 years | 2 |
| 10-17 years | 3 |
| 18-30 years | 2 |
| >30 years | 2 |
| **Sex** |  |
| Female | 4 |
| Male | 5 |
| **General Symptoms** |  |
| Progressive weight loss | 7/8* |
| BMI 17 to <21 kg/m<sup>2</sup> | 2 |
| BMI 15 to <17 kg/m<sup>2</sup> | 2 |
| BMI <15 kg/m<sup>2</sup> | 5 |
| Fatigue | 4 |
| Muscle weakness | 6 |
| **Respiratory Symptoms and findings** |  |
| Progressive exertional dyspnea | 9 |
| Dyspnea at rest | 8 |
| Chronic cough | 5 |
| Deepened suprasternal notch | 8 |
| Restrictive ventilatory defect | 9 |
| FVC <40% predicted | 5/5* |
| Hypoxemia at rest while breathing room air | 1/6* |
| **Lung Biopsy findings** |  |
| Lung Biopsy performed | 4 |
| Histologically confirmed subpleural alveolar fibroelastosis | 4 |
| **Chest CT Features** |  |
| Findings predominantly in the upper lobes | 9 |
| Pleural and septal thickening | 9 |
| Subpleural reticular opacities | 8 |
| Traction bronchiectasis | 4 |
| Patchy ground glass opacifications | 7 |
| Honeycombing | 0 |
| Pneumothorax ever | 6 |
| Platythorax | 5 |
| Tracheal deviation | 7 |
| **Extrapulmonary Organ Manifestation** |  |
| Pulmonary hypertension | 1 |
| Elevated liver enzymes with normal synthesis function | 5 |
| After liver transplantation | 2 |
| Proteinuria/Nephropathy | 4 |
| Peripheral polyneuropathy | 5 |
| Neurodevelopmental delay | 2 |
| **Status at last Follow-up** |  |
| Alive (one patient post lung transplantation) | 7 |
| Death | 2 |

Data was collected during the initial presentation. The term BMI denotes Body mass index, FVC Forced vital capacity, DLCO Diffusion capacity of the lung for carbon monoxide, CT Computerized tomography. Numbers with asterisks above indicate that information was not available for all patients.

### Germline biallelic loss-of-function variants in *DGUOK*

We performed whole exome or whole genome sequencing in nine patients and identified biallelic germline variants in *DGUOK*. In F1 (P1, P2), F2 (P4), and F4 (P6-P9) the same intronic homozygous variant (c.592-26A>G p.(Val198Alafs*4)) was detected (ENST00000264093.9) (Figure 2A, exemplified for P1 and P2). P3 (F1) carries this intronic variant in compound heterozygosity with a nonsense variant (c.313C>T p.(Arg105*)) and P5 (F3) has a homozygous four-base-pair duplication (c.763_766dup p.(Phe256*)). Therefore, the clinical phenotype of P1, P2, P4, P6-P9 differs from that of P3 and P5 with no liver involvement and later onset of respiratory symptoms (Clinical phenotype and Table S3, Supplementary Appendix). All tested healthy relatives were either heterozygous carriers of one of the familial variants or wild type (Figure 1).

**Figure 2.**
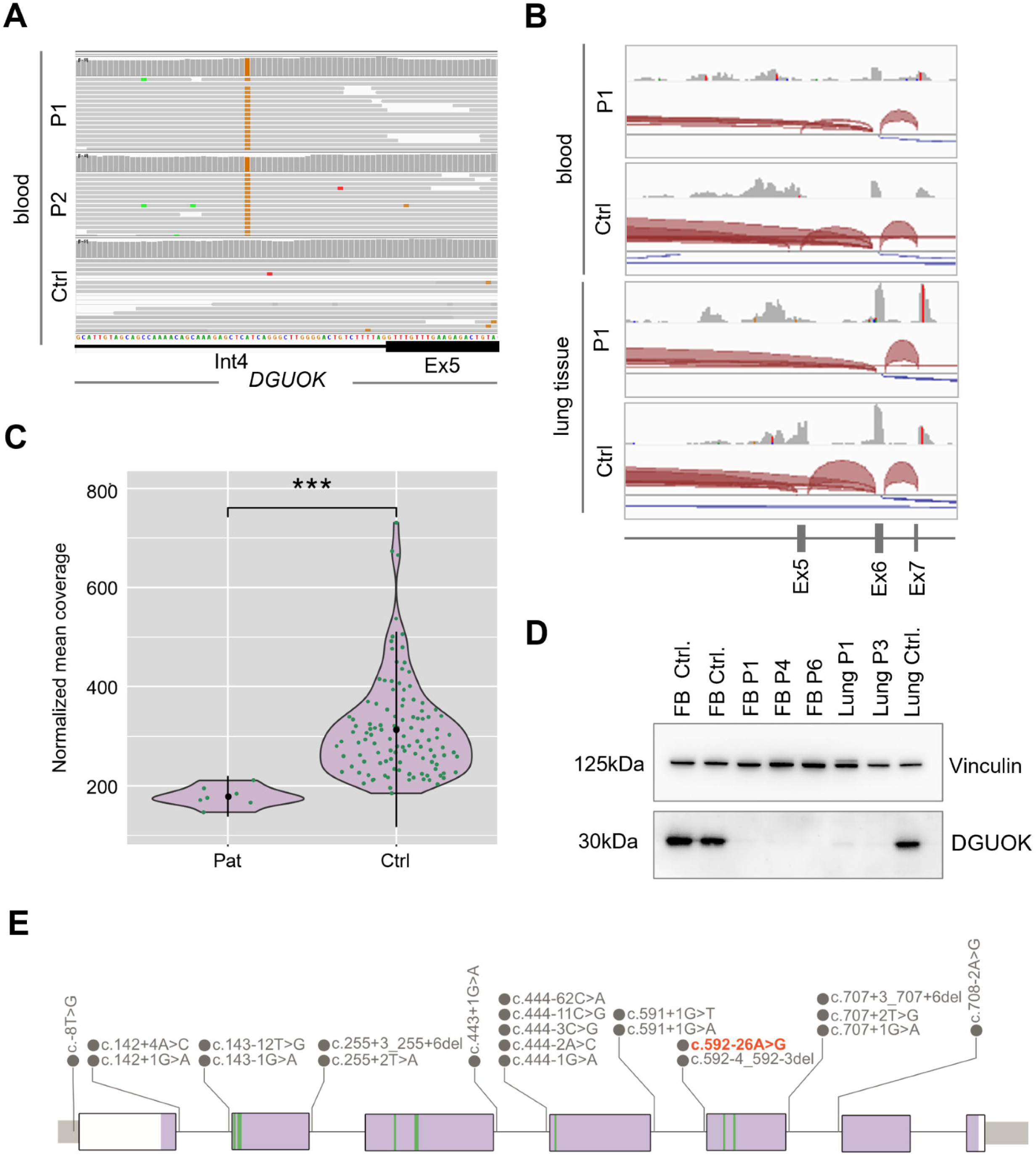
Genomic and transcriptomic analysis of *DGUOK* locus. Panel A shows an Integrative Genomics Viewer (IGV) screenshot from WGS analysis of peripheral blood from P1 and P2 highlighting an adenine-to-guanine substitution present in all reads (homozygous occurrence), located 26 base pairs upstream of the intron–exon boundary between intron 4 and exon 5. The control sample (Ctrl) shows no deviation from the reference sequence at this position. Panel B depicts RNA-seq read coverage tracks for blood and lung tissue of P1 and a control (Ctrl). Red arcs indicate splice junctions. The exon–intron structure of the gene is displayed at the bottom, with exons 5–7 indicated. In P1 the variant c.592-26A>G in *DGUOK* leads to exon 5 skipping in the majority of blood-derived RNA reads compared to Ctrl, while lung-derived RNA of P1 shows exclusive exon 5 skipping. Panel C illustrates the amount of mitochondrial DNA (mtDNA) in patients carrying the homozygous variant c.592-26A>G in *DGUOK* (Pat) and a control group (Ctrl). Each point represents an individual sample, and the distribution width indicates the density of the data. The normalized mean coverage of mtDNA is significantly higher in the control group compared to the case group, as indicated by the triple asterisk (***), denoting a highly significant difference (*p* < 0.001). The black bars represent the median and interquartile range. Panel D represents western blot showing DGUOK expression in control (Ctrl) and patient’s material (Fibroblasts (FB) of P1, P4, P6, Lung of P1, P3). Vinculin served as a loading control. Panel E represents the *DGUOK* gene structure with previously reported intronic and splice-region variants above the gene model. Exons are shown as purple boxes and introns as connecting lines. The intronic variant c.592-26A>G, identified in this study, is highlighted in red. Green bars indicate substrate-binding sites.

The variants c.313C>T p.(Arg105*) and c.763_766dup p.(Phe256*) are predicted to lead to premature stop codons with subsequent mRNA degradation via nonsense-mediated decay (NMD), while the intronic c.592-26A>G variant was predicted to cause aberrant splicing based upon the computer-based splicing site prediction program spliceAI score of 0.5 and a branch point predicted value of 5.4. Therefore, sequencing was performed on blood-derived RNA (P2) and RNA isolated from explanted lung tissue obtained after LTx (P1). The red splice junction reads shown in Figure 2B indicate that exon 5 skipping occurs in many, but not all, blood-derived transcripts. In contrast, in lung tissue this variant results in skipping of exon 5 in all transcripts. Via Sanger cDNA sequencing exon 5 skipping was shown to cause a frameshift (data not shown), which most likely leads to the introduction of a premature stop codon in exon 6 (p.(Val198Alafs*4)), potentially resulting in degradation of the affected transcripts via NMD. At the protein level, DGUOK expression was absent or markedly reduced in all patient-derived samples (1D). Several intronic (splice-) variants have been identified so far in patients with DGUOK-deficiency (Figure 1E) but none of them appears to result in the phenotype described here, most likely causing different effects at the protein level compared to those observed in our patients.

### Reduced blood mtDNA and tissue-specific respiratory chain dysfunction

The quantification of the average mitochondrial DNA (mtDNA) revealed a clear reduction in mtDNA levels in the blood of patients carrying the homozygous variant, consistent with a mitochondrial DNA depletion syndrome (Figure 2C). To determine the effect of the mitochondrial DNA depletion on mitochondrial oxidative phosphorylation (OXPHOS) in patient-derived fibroblasts (P1, P3, P4), we analyzed respiratory chain enzyme activities and performed mitochondrial stress tests to assess oxygen consumption rates (OCR). Analysis of respiratory chain enzyme activities revealed values within the normal reference ranges for complexes I+III, IV (COX), and V (ATP synthase). The activities of complexes II+III were at the lower limit of the reference interval or slightly below it (P4). Consistently, Seahorse mitochondrial stress testing showed no statistically significant differences in maximal OCR between patient-derived fibroblasts and controls, despite a trend toward higher OCR in patient cells (Figure S1, Supplementary Appendix). Together, these findings indicate preserved mitochondrial respiratory chain function in the analyzed fibroblasts. However, analysis of muscle tissue of patient P3 revealed a mitochondrial defect. In the muscle, activities of complexes II, III, and IV were markedly reduced, with complete loss of cytochrome c oxidase (COX) activity, despite normal citrate synthase activity, indicating preserved mitochondrial content. These findings demonstrate a tissue-specific manifestation of mitochondrial respiratory chain dysfunction (Figure S1, Supplementary Appendix).

### Emergence of Aberrant Basaloid Cells in patients with PPFE and DGUOK-associated mitochondriopathy

To assess the cellular and transcriptional impact of *DGUOK* dysfunction in the human lung, we performed single nucleus RNA sequencing (snRNAseq) on six lung samples from four pediatric patients (P1-P4). In total, 66947 single epithelial nuclear transcriptomes (with 26120 from patients with DGUOK deficiency) were analyzed, enabling identification of all principal lung epithelial cell types and assessment of cell frequencies across disease and control samples (Figure 3A-B). We identified a population lacking basal (*KRT5, KRT15*), alveolar type I (AT1) cells (*AGER*), and AT2 (*SFTPC, SFTPA, LAMP3*) markers, while expressing mesenchymal and cell–matrix–associated genes (*ITGAV, ITGB6, CDH2*), injury- (*CTSE, PTGS2)* and senescence-associated genes (*CDKN2B, GDF15*). Expression of additional epithelial lineage–associated markers (*KRT17, TP63*) confirmed these cells as Aberrant Basaloid cells^27^ (Figure 3E–G; Figure S2 and Table S4, Supplementary Appendix). These cells arise from injured alveolar type II (AT2) cells via intermediate cells (iAT2), which we likewise identified (Figures 3 and 4A–D). iAT2 cells express some overlapping AT2 and Aberrant Basaloid markers with the addition of injury-response genes (*CTSE, MYO16, NDNF*). iAT2 and Aberrant Basaloid cells frequencies were significantly increased in patients with DGUOK deficiency, representing 8.3% (vs 0.9 % in controls, adjusted p<0.005) and 6.1 % (vs 0.5 % in controls, adjusted p<0.002) of epithelial cells, respectively (Figure 3C).

**Figure 3.**
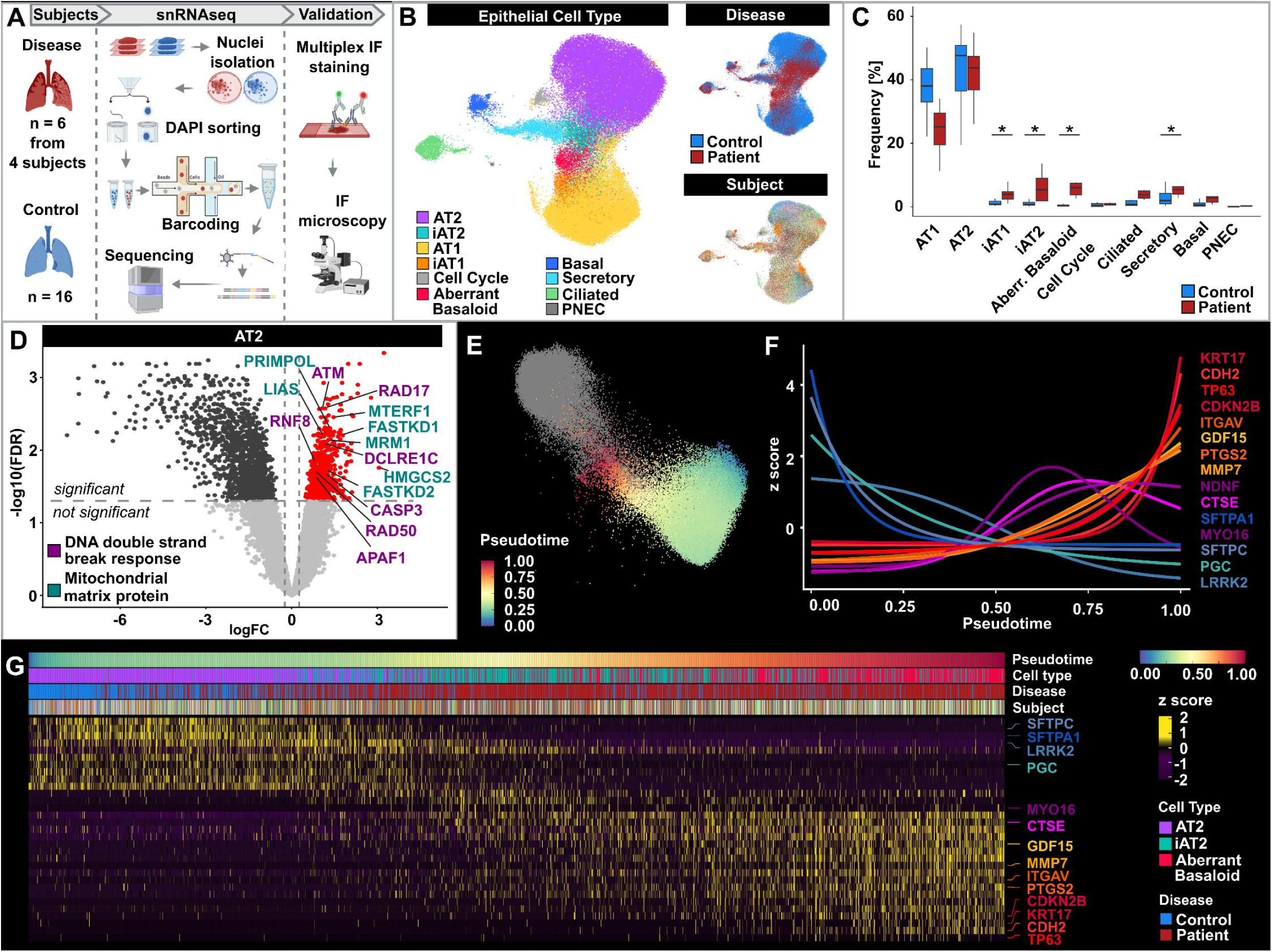
snRNAseq reveals AT2 transition to Aberrant Basaloid cells in DGUOK-associated PPFE. Panel A schematically illustrates the steps of the snRNAseq experiments. Six lung tissue samples from four patients were analyzed. Lung tissue samples from 16 subjects with a physiologic appearance served as controls. Nuclei were isolated and sorted by DAPI-based cell sorting, then encapsulated in droplets with barcoded beads. Within each droplet, transcripts were labeled with cell-specific barcodes and unique molecular identifiers (UMIs), and the resulting libraries were sequenced. Cell populations of interest were localized by multiplex immune fluorescence (IF) staining and IF microscopy. Panel B shows three Uniform Manifold Approximation and Projections (UMAPs) of epithelial cells, annotated by cell type, disease, and subject. Panel C shows significant differences in the relative frequencies of epithelial cell types between patients with PPFE and DGUOK-associated mitochondriopathy and controls, including the presence of intermediate cells (iAT2) and Aberrant Basaloid cells (Aberr. Basaloid). Box plots show the median, interquartile range, and full range of values, Asterisks indicate statistical significance (*p<0.05). Panel D shows a volcano plot of changes in gene expression between control and disease, with positive values indicating higher expression in disease. Genes involved in mitochondrial matrix proteins (green) and in DNA double strand break response (purple) show significantly increased expression. Panels E depicts the pseudotime trajectory from AT2 via intermediate cells (iAT2) to Aberrant Basaloid cells. In Panel F, a generalized additive model illustrates the genes changing from AT2 to Aberrant Basaloid cells as a function of pseudotime, while Panel G shows a heatmap of the corresponding genes, with each column representing a single nucleus randomly sampled from fixed intervals.

**Figure 4.**
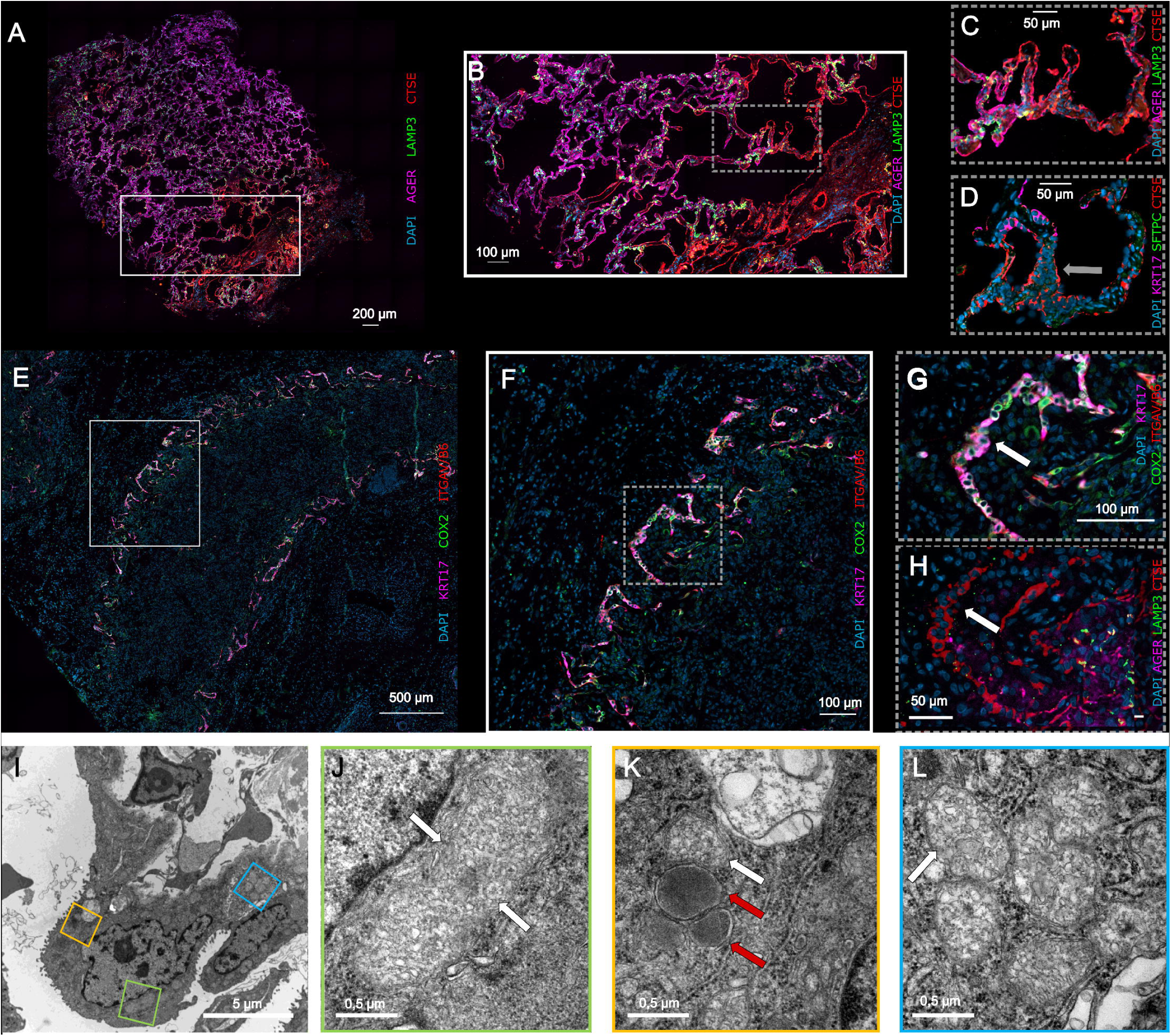
Aberrant Basaloid cells are localized at the fibro-alveolar remodeling edge. Panel A to H show representative immunofluorescence stains of lung tissue from two pediatric patients (Panel A-D: cryobiopsy of P4; Panel E-H: lung explant of P1) with PPFE and DGUOK-associated mitochondriopathy. Intermediate cells (iAT2) as precursor cells of Aberrant Basaloid cells are found expressing CTSE but not KRT17, as indicated by the gray arrow in Panel D. Aberrant Basaloid cells localize at the fibro-alveolar remodeling edge, highlighted by white arrows in Panels G and H. They were characterized by positivity for CTSE, COX2, ITGAV/ITGB6, and KRT17, and by absence of expression of AGER, a specific marker for AT1 cells, and SFTPC and LAMP3, a specific marker for AT2 cells. Panel A, B, C and H show LAMP3 (green), CTSE (red), AGER (pink). Panel D depicts SFTPC (green), CTSE (red) and KRT17 (pink). Panel E, F and G demonstrate COX2 (green), ITGAV/B6 (red) and KRT17 (pink). Nuclei are stained with DAPI. Panel I shows an overview of the ultrastructure of an Aberrant Basaloid cell and panels J to L some details with a focus on the mitochondria (white arrows). These are enlarged and characterized by a loss of mitochondrial matrix as well as swelling and lysis/ degeneration of their cristae (Panels J-L). The red arrows in Panel K highlight electron dense bodies as a typical feature of Aberrant Basaloid cells.

Comparing gene expression across epithelial cell types, AT2 cells exhibited the strongest differential gene expression, with over 2,000 genes significantly altered between patients and controls. Specifically, nuclear-encoded genes involved in DNA double-strand break response (*ATM*, *RAD50*, *RNF8*) as well as mitochondrial matrix proteins (*MTERF1*, *FASTKD2*, *FASTKD1, PRIMPOL*), were upregulated (Figure 3D, Table S5, Supplementary Appendix).

Additional alterations were observed across endothelial, mesenchymal, myeloid, and lymphoid compartments (Figures S3 and S4 and Table S5, Supplementary Appendix). In particular, a relative expansion of profibrotic and elastofibrotic fibroblasts was detected, along with increased frequencies of regulatory T and B cells and reduction of T Helper cells, collectively shaping the profibrotic microenvironment.

Immunofluorescence stains confirmed the presence of Aberrant Basaloid cells, which form a monolayer at the fibrotic edge, surrounded by iAT2 cells (Figure 4A-H).

In view of the changes in gene expression and the emergence of Aberrant Basaloid cells in lung tissue from our patients, we anticipated corresponding mitochondrial alterations. Thus, electron microscopy of lung tissue was performed to assess mitochondrial ultrastructure. In Aberrant Basaloid cells, electron microscopy revealed numerous, enlarged mitochondria characterized by a loss of mitochondrial matrix as well as swelling and degeneration of their cristae (Figure 4I-L). Degenerating mitochondria were already evident in AT2 cells (Figure S5, Supplementary Appendix), whereas no alterations were observed in skin fibroblasts (Figure S6, Supplementary Appendix), underscoring the tissue-selective impact of DGUOK deficiency.

## Discussion

This study highlights several novel key aspects: We present the first pulmonary phenotype of a mitochondrial depletion syndrome. We demonstrate that a homozygous intronic variant in *DGUOK* leads to a clinically distinct pulmonary and later onset phenotype, compared to the classical early-onset hepatocerebral phenotype in patients with DGUOK-associated mitochondriopathy. More specifically, DGUOK dysfunction represents the first monogenic, non-telomeric cause of PPFE and therefore justifies the definition of hereditary PPFE (hPPFE). Our study suggests that mitochondrial dysfunction plays a pivotal role in the pathogenesis of PPFE, although energy metabolism in lung cells was not examined. Last, this is the first description of Aberrant Basaloid cells in pediatric patients linking pediatric mitochondrial disease to adult fibrosing lung pathology.

In our cohort, seven patients carrying the intronic homozygous hypomorphic c.592-26A>G variant exhibited no evidence of severe neonatal, infantile, or late-onset hepatocerebral involvement^23,25^ and remained symptom-free at least until adolescence. All patients presented with respiratory symptoms, weight loss, and peripheral polyneuropathy as the predominant manifestations at disease onset. Highly likely, this phenotype results from partial preservation of *DGUOK* function, allowing residual production of functional protein, which may be sufficient to maintain hepatic function in early childhood. Over time, however, chronically reduced DGUOK levels may lead to progressive involvement of other organ systems, particularly the lung.

To date, detailed analyses of lung disease have not been reported in patients with DGUOK deficiency or in any other hereditary mitochondrial disorder^21,23^. However, as mentioned above, respiratory involvement was reported in 13% of 173 subjects in a large cohort of individuals with DGUOK deficiency, but a pulmonary phenotype was not further characterized^23^. A retrospective analysis of a chest CT from patient P5 with a homozygous frameshift variant, who underwent neonatal liver transplantation, revealed a previously unrecognized PPFE pattern that ultimately accounted for the death of the patient. Similarly, P3 recently died from progressive respiratory failure. In a retrospective analysis of hepatic manifestations in mtDNA depletion syndromes, Vara et al. (2023) reported one patient harboring the intronic variants c.592-26A>G and c.444−3C>G, who presented with early-onset liver disease and an unspecified interstitial lung disease^24^. This report and our findings in P3 and P5 suggest that severe pulmonary involvement is not restricted to hypomorphic variants, and that PPFE may have been overshadowed by the severe liver and neurological involvement or was overlooked in patients with pulmonary hypertension so far, which is described as a significant complication in some patients with DGUOK deficiency after liver transplantation^21,23^.

Although a definitive genotype–phenotype correlation has not been established in DGUOK deficiency and substantial inter- and intrafamilial variability exists with respect to both the type and severity of clinical manifestations, the available data suggest that earlier disease onset is generally associated with a more severe loss of gene function, whereas late-onset phenotypes are more likely to reflect residual enzymatic activity. In the largest published summary of reported cases, Manzoni et al. reported genotype–phenotype data for 202 patients^21^. Among 88 patients presenting with neonatal or infantile hepatomyocerebral, hepatocerebral, or isolated hepatic disease, 66 carried at least one truncating or splice-site variant. The remaining 22 patients harbored biallelic missense variants, of which more than half involved a glutamate-to-lysine substitution, likely impacting protein structure, stability, and function.

We provide the first detailed cellular characterization of pulmonary involvement in a defined mtDNA depletion syndrome, identifying Aberrant Basaloid cells - a pathological cell population previously described in adult, age-associated fibrosing lung diseases - in a pediatric cohort. Collectively, this reframes mitochondrial dysfunction as a central driver of PPFE pathogenesis, rather than an acquired contributor. Disruption of the mitochondrial membrane, inhibition of the respiratory chain, and reduced enzyme activity^28^, all reflected by abnormally enlarged and swollen mitochondria with lysis or degeneration of cristae in the ultrastructure, may trigger cellular damage processes and increased production of reactive oxygen species (ROS). snRNAseq revealed increased expression of nuclear-encoded genes involved in DNA double-strand break response and mitochondrial transcription in AT2 cells, consistent with DGUOK-related mitochondrial stress associated with impaired mtDNA maintenance and increased ROS accumulation.

Once established at the fibrotic edge, Aberrant Basaloid cells sustain fibrotic remodeling by engaging profibrotic (myo-) fibroblasts, thereby reinforcing the usual fibrotic niche^27,29^ and contributing to iPPFE development and rapid disease progression.

Our findings have several clinical implications: First, as PPFE in DGUOK deficiency manifests clinically much later than classic forms, and presents as a phenotype distinct from other mitochondrial disorders, DGUOK deficiency and other mitochondriopathies should be considered in patients with typical features of presumed PPFE, irrespective of age at onset and even in the absence of typical mitochondrial features. Hence, genetic testing may be considered in the diagnostic evaluation of patients with suspected iPPFE.

Second, once symptomatic, disease progression in our patients was rapid, with some developing end stage lung disease or dying from respiratory failure within a short time after diagnosis. It can be assumed that the disease manifests subclinically much earlier, but patients only develop symptoms at an advanced stage because gas exchange is maintained for a long time due to a limited number of affected alveoli^4^. For example, P2 had only mild pulmonary complaints and was functionally well at the time of diagnosis, although the patient already had a severely impaired lung function and typical radiological and histological findings of PPFE. These observations emphasize the importance of early recognition of PPFE-specific clinical and radiologic features, careful longitudinal monitoring, and tailored management strategies. Third, we provide evidence for a hereditary component of PPFE, previously suggested by familiar cases with affected siblings^33^, although a definitive genetic cause has remained elusive^6,8–11^. With this report, we propose expanding the classification of pleuroparenchymal fibroelastosis to include hereditary forms (hPPFE).

Fourth, as there are currently no effective drugs available for DGUOK deficiency, lung transplantation is the only treatment option in some but not all patients. Therefore, there is an urgent need for the development of novel pharmacologic therapies. Future research approaches include nucleotide replacement therapy with dGMP and dAMP, which might partially support mitochondrial DNA synthesis, and second, antisense oligonucleotide (ASO)–based approaches, which could potentially modulate or correct some *DGUOK* variants. Fifths, our findings raise the possibility that previously presumed isolated pulmonary hypertension after liver transplantation in DGUOK patients may in fact be secondary to PPFE, as pulmonary hypertension has been reported in approximately 20% of PPFE cases^30^.

In summary, our findings highlight that mitochondrial DNA depletion syndromes can manifest as primary fibrosing lung disease and suggest that pulmonary involvement represents an underrecognized feature of the mtDNA depletion syndrome-spectrum, extending beyond the traditional early-onset phenotypes. Furthermore, our findings underscore the necessity of comprehensive molecular analysis and genetic counseling in cases of unexplained fibrotic lung disease.

## Data Availability

All data produced in the present study are available upon reasonable request to the authors.

## Acknowledgements

We would like to thank the families for their participation. We thank Dr. Jens Dingemann, Dr. Julia Brendel and Dr. Fabio Ius for their dedicated surgical care of the patients. We thank Duygu Halibryam for her Turkish translation and assistance. We thank Caterina Garone and Peter Freisinger for the constructive discussion on DGUOK deficiency. We thank Andreas Schmiedl for his assistance in electron microscopy. We acknowledge that AI-assisted language tools were used solely for grammar and style improvements and did not influence the scientific content or data interpretation.

## Contributors

SvH, NS, JS and PM contributed to the study conception and design.

Experiments and data collection were performed by SvH, NS, JCS, PM, AMD, DJ, AH, ADu, TH, LC

Patient recruitment, clinical assessment and sample collection were conducted by NS, JCS, PM, DS, BS, NJ, LH

WES, WGS, snRNAseq analysis, bioinformatics and/or data analysis were performed by SvH, NS, JCS, PM, LK, MS, DJ, KP, JR, AS, WH, BS

Supervision was provided by NDD, BA

The first draft of the manuscript was written by SvH, NS, JCS, PM, LK All authors reviewed and approved the manuscript.

## Data sharing

The datasets generated and analyzed for this study are not publicly available due to concerns regarding participant/patient anonymity. Requests to access the datasets should be directed to the corresponding author.

