## Supplementary material for "Pleuroparenchymal fibroelastosis in monogenic *DGUOK*-associated mitochondriopathy": Supplementary Appendix.pdf

### I. Supplementary methods

#### Patient cohort, ethics approval, and consent to participate

Nine patients with genetically confirmed DGUOK-deficiency (five children and four adults) have been included. Data on symptoms and clinical courses were obtained from clinical records and treating physicians. The study adhered to the principles of the Declaration of Helsinki. Patients provided assent, and their parents provided informed consent for the use of data and biomaterials, either through inclusion in the European Registry for childhood interstitial lung disease (chILD-EU Registry), via the Broad Consent of the German Center for Lung Research (DZL), or as part of the treatment contract of Hannover Medical School. DZL Broad consent and enrollment in chILD-EU registry was approved by the local ethics committee of the Hannover Medical School, Germany (Ethical vote 2923-2015, 1969-2013 and 12241-BO-K-2026, respectively).

#### Whole exome or genome sequencing and variant interpretation

Whole-exome sequencing (WES) (P4, P5) or whole-genome sequencing (WGS) (P1, P2, P3, P6-P9) was performed at Hannover Medical School on DNA extracted from peripheral blood samples collected in ethylenediaminetetraacetic acid (EDTA) tubes. DNA extraction, library preparation, and sequencing were conducted using standard protocols. Bioinformatic analyses, including alignment, variant calling, annotation, filtering, prioritization, and evaluation, were performed using our center-specific pipeline. Sequencing reads were aligned to the human reference genome (GRCh38), and variant calling was conducted according to established best practices. Variant interpretation incorporated allele frequency data and computational prediction tools, including phyloP, SIFT, PolyPhen-2, FATHMM, CADD, and REVEL. Identified variants were further assessed using public databases, including the Leiden Open Variation Database ClinVar (https://www.ncbi.nlm.nih.gov/clinvar), and the Genome Aggregation Database (gnomAD; https://gnomad.broadinstitute.org/). The classification of sequence variants was performed in accordance with the standards and guidelines of the American College of Medical Genetics and Genomics (ACMG)[31].

#### Whole blood RNA sequencing analysis

Whole blood collected in PAXgene Blood RNA tubes (PreAnalytics GmbH, Switzerland) and lung tissue preserved in RNAlater (ThermoFisher) were used for whole-transcriptome analysis. Total RNA was extracted according to the respective manufacturers' protocols. RNA quality and quantity were assessed by spectrophotometry. RNA libraries were prepared for sequencing using Takara SMART-Seq stranded total RNA Kit and processed on an Illumina NovaseqXplus platform with standard paired-end runs. Downstream data analysis included alignment to the GRCh19 reference genome, expression quantification, and pathway enrichment analysis using established bioinformatic pipelines (STAR version 2.7, Center for Human Genetics, Germany, Tübingen). Sashimi plots and coverage profiles were generated using the IGV Genome Browser v2.18.1.

### Cell culture

3 mm punch biopsies of patient P1, P3 and P4 as well as the healthy mother of P1/ P2 and the healthy father of P4 were obtained under local anesthesia and transported in sterile PBS or cell culture media with antibiotics. The tissue was minced under sterile conditions and cultured in DMEM with 10% fetal bovine serum (Sigma-Aldrich), 1 % sodium pyruvate (Gibco) and 1 % penicillin-streptomycin (Gibco) at 37°C with 5% $CO_2$. Low passages of fibroblasts cultures migrating from the explant were expanded and used for further experiments.

### Western Blot Analysis

Protein lysates were prepared from fibroblasts using the RIPA lysis buffer system (Santa Cruz). After electrophoresis, proteins were transferred onto an Amersham Protran nitrocellulose membrane (Sigma/Merck, Darmstadt, Germany). The membrane was blocked for 1 h in non-fat milk and incubated with primary polyclonal antibody (anti-DGUOK Invitrogen Immunogen PA5-63419, anti-Vinculin #18799 Cell signaling) overnight at 4°C and secondary antibody (Goat anti rabbit IgG-HRP #7074 Cell signaling, Goat anti rabbit IgG-HRP, abcam #ab6721) at room temperature for 2 h, respectively.

### Respiratory chain enzyme analysis in fibroblasts and muscle tissue

Muscle biopsies of P3 for respiratory chain enzyme analysis were snap-frozen immediately after collection by plunging the fresh tissue into liquid nitrogen and stored at –80°C to preserve mitochondrial enzyme activity. Fibroblasts of P1, P3 and P4 were cultured in T25 flask and then plated on Petri dishes with a diameter of 2 cm. Fibroblasts were sonicated to expose respiratory chain enzymes. Respiratory chain complexes (complex I+III, complex II+III, cytochrome c oxidase and ATP-synthase as well as citrate synthase) were assayed as described before[1]. A postnuclear supernatant of the skeletal muscle biopsy was prepared and ATP-synthase assayed as described previously[2]. Complex I+III, complex II+III and cytochrome c-oxidase as well as citrate synthase enzyme activities were measured using a slight modification of the methods described above for fibroblasts.

**Mitochondrial DNA content calculation**

The calculation of the mean coverage of the mitochondrial chromosome and the autosomes of each sample was performed with PanDepth[3] by using the Whole genome sequencing (WGS)-cram files of the seven patients with the same homozygous pathogenic variant (c.592-26A>G, P1, P2, P4, P6-P9) and of 115 randomly chosen controls. WGS was performed on blood samples from the respective individuals. Mitochondrial DNA content calculation was done according to the formula 2x(chrM coverage)/(genome coverage) [4] using bc (https://www.gnu.org/software/bc/). Visualization was done with the ggplot2 and ggsignif packages on R version 4.5.2.

**Transcriptomic Analyses**

Nuclei isolation from FFPE tissue for Chromium Fixed RNA Profiling

From each formalin-fixed paraffin-embedded (FFPE) tissue block, three sections with a thickness of 50 µm were prepared and placed into Miltenyi C-tubes. Deparaffinization was performed at room temperature by immersing each specimen in xylene for three consecutive 10-minute treatments. For each specimen and wash, 1 mL of xylene was used. Tissue was rehydrated through an ethanol series using 100% ethanol, 70% ethanol, 50% ethanol, and 30% ethanol. Post rehydration, each specimen was washed thrice with 1X phosphate-buffered saline (PBS) containing 0.5 mM calcium chloride. The Tissue Digestion Buffer (TDB, see table Y) was pre-warmed at 37°C for 10 minutes before use. Two milliliters of TDB were added to each specimen in C-tubes, before loading onto a gentleMACS Octo Dissociator and processed using the following program: (1) incubation at 37°C for 45 minutes; (2) clockwise rotation at 2,000 rpm for 30 seconds at 37°C; (3) counterclockwise rotation at 2,000 rpm for 30 seconds at 37°C; and (4) removal of the C-tubes. Finally, the C-tubes were briefly centrifuged at 300 × g for approximately 30 seconds to collect all nuclei at the bottom of the tube, before resuspension in the supernatant. The dissociated tissues were filtered through 70 µm mesh filters (PluriSelect #43-50070-51) to remove debris and undissociated tissue fragments. To maximize recovery, each C-tube was first rinsed with 1 mL of 1X PBS, which was passed through the filter, and the filter itself was subsequently rinsed with an additional 1 mL of 1X PBS. Filtrates from both steps were collected in the same tube as the corresponding dissociated tissue. The specimens were centrifuged at 850 × g for 5 minutes at 4°C, and supernatants were carefully removed without disturbing the pellets. Each pellet was resuspended in 1 mL of chilled Quenching Buffer (content of 10X Genomics, #1000781) and subsequently passed through a 20 µm filter (PluriSelect #43-10020-50) to remove debris and any remaining undissociated cells. Nuclei quality and count was assessed using a LUNA-FL™ Dual Fluorescence Cell Counter with the Acridine Orange/Propidium Iodide (AO/PI) Cell Viability Kit (BioCat, #F23001-LG). Nuclei suspensions were gently mixed by pipetting, and 18 µL of per suspension was combined with 2 µL of AO/PI. LUNA™ reusable slides were loaded with 10 µL of this mixture per chamber. All nuclei suspensions were stored overnight at 4°C with 10% enhancer (content of 10X Genomics, #1000781) prior to Flow-Activated Nuclei Sorting (FANS): Enhancer was thawed at 65°C for 10 minutes, briefly vortexed, and spun down. Before use, it was kept warm and inspected to ensure no precipitate was present. Pre-warmed Enhancer was then added to each nuclei suspension at a ratio of 1:10 and gently mixed by pipetting.

**Supplemental Table S1: Buffers used for snRNA-sequencing:**

| Buffer | Compound | Manufacturer (Catalog Number) | Stock concentration | Final concentration |
|---|---|---|---|---|
| TDB | Liberase™ TM Research Grade | Merck/ Roche (5401127001) | Lyophilized, prepared as 1 mg/mL | 250 µg/mL |
| | Collagenase D from Clostridium histolyticum | Merck/ Roche (11088882001) | Lyophilized, prepared as 100 mg/mL | 2.5 mg/mL |
| | Calcium Chloride Solution | Jena Bioscience (BU-103) | 2.5 M | 0.5 mM |
| | PBS - Phosphate-Buffered Saline | Invitrogen™ (AM9624) | 10X | 1X |
| | Protector RNase-Inhibitor 10 000 U | Merck/ Roche (3335402001) | 40 U/µL | 0.04 U/µL |
| | Nuclease-free H2O | Invitrogen™ (AM9932) | N/A | To 2 mL |

**Isolation of DAPI$^+$ Single Nuclei by Fluorescence-Activated Nuclei Sorting (FANS)**

Immediately prior to sorting, nuclei suspensions were stained with 4′,6-diamidino-2-phenylindole (DAPI, Thermo Scientific™ #62248), with final concentrations ranging from 1 to 10 µg/mL and adjusted as needed to achieve optimal staining. Sorting was carried out using cell sorters equipped with a 100 µm nozzle at 35 psi (FACSAria III Fusion, FACSAria IIu, Becton-Dickinson, or MoFlo XDP, Beckman-Coulter) in the central research facility Cell Sorting of Hannover Medical School. For each specimen, 5 × $10^5$ DAPI$^+$ single nuclei were collected into individual 15-mL tubes, each containing 1 mL of 0.5X PBS with 0.02% BSA (Sigma-Aldrich, #126615-25ML). Intact single nuclei were isolated by flow cytometry based on light scattering characteristics and DNA content, as indicated by DAPI fluorescence. A tight gate was applied in the DAPI channel to exclude nuclei with suboptimal DNA content and debris. Doublets were removed in subsequent gating steps using scatter pulse parameters, and the main population was further refined using an FSC-SSC gate. Reanalysis of the nuclei from each specimen confirmed a homogeneous population. Most specimens yielded the desired 300,000 nuclei, besides one sample of patient 2 and one of 3, of which all sorted nuclei were used for further analysis. The sorted single nuclei suspensions were kept on ice until processing with the Chromium Fixed RNA Profiling Reagent Kits for Multiplexed Samples.

**Chromium Fixed RNA Profiling of Multiplexed Samples**

Sorted DAPI$^+$ single nuclei were immediately processed following the User Guide CG000527, Rev B (Chromium Fixed RNA Profiling Reagent Kits for Multiplexed Samples), as briefly summarized below. Hybridization was performed in PCR tube-strips at 42°C for 19h. Due to exact determination of nuclei count, samples were pooled after hybridization for a pooled wash in Post-Hyb Wash Buffer B (contains 10X Genomics, #2001308 and #2000482). After the third rinsing step, samples were resuspended in 100 µL and filtered through a 20 µm filter. Nuclei concentration was determined thrice, with 2 µL of sample pool, 16 µL of ddH2O and 2 µL of AO/PI of which 10 µL were counted. GEM-X FX Chip was loaded according to instructions with a target nuclei recovery of 320,000. The chip was loaded into the Chromium X and GEMs were transferred immediately after instrument run. GEM incubation, GEM recovery and pre-amplification were performed on the same day, DNA cleanup and library preparation the next day. cDNA libraries were stored at -20°C until sequencing.

**Sequencing & Data processing of snRNAseq data**

Libraries were sequenced on an Illumina NovaSeqX platform, with a sequencing configuration of 28 base pairs for Read1, 90 base pairs for Read2, and 10 base pairs for Index5 and Index7. Then, base calls were converted to reads using the Cell Ranger software (8.0.1) and its bcl2fastq implementation ("mkfastq"). Reads were aligned to the human reference genome GRCh38 (GENCODE v32/Ensembl 98, "GRCh38-2020-A") and the Chromium Human Transcriptome Probe Set _v1.0.1 for GRCh38-2020-A. Count matrices were analyzed using the Seurat pipeline (R package Seurat v5.4.0 in R v4.4.3), following recommendations for scRNAseq data processing and integration https://satijalab.org/seurat/). The newly generated data from 6 samples from 4 DGUOK patients, 2 pediatric control and one adult control subjects was merged with 13 previously generated high quality control samples, generated with the same technology (GSE284081, GSE301982). Low quality filtering was conducted in an iterative adaptive fashion per sample. Nuclei with > 5% mitochondrial counts, < 300 total counts, and < 200 features were filtered out prior to downstream analysis. UMI counts were normalized by applying a scale factor of 10,000 UMIs per cell. This was followed by natural log transformation with a pseudocount of 1 in accordance with methods previously published by our group and others. UMI counts were normalized using a scale factor of 10,000 UMIs per cell, followed by natural log transformation with a pseudocount of 1. The most variable genes were identified using Seurat's FindVariableGenes (nFeatures = 3000) function. Integration was performed using IntegrateLayers (method = scVIIntegration).

### Integration, dimensionality reduction, clustering, cell type annotation of snRNAseq data

Louvain clustering was then performed, and for visualization, nuclei and their graph-based embeddings were projected into a low-dimensional space using Uniform Manifold Approximation and Projection (UMAP). Clusters lacking differential gene expression were merged. Cell lineages (i.e., "Epithelial," "Stromal", "Immune") were then assigned based on lineage-specific marker gene expression. This procedure was repeated twice for each subset, incorporating graph embedding and cluster analysis to identify multiplets and cellular debris, which were removed prior to downstream analyses. The cleaned lineage-specific datasets were subsequently integrated using scVI integration, followed by final dimensionality reduction and graph embedding as described above. Cell types were identified through cluster-specific marker genes as determined by the Wilcoxon rank-sum tests implemented in Seurat's FindMarkers function, comparing all cells within each cluster to all other cells in the dataset. To account for multiple testing, p-values were adjusted using Bonferroni correction method, with adjusted p-values < 0.05 considered statistically significant. Final cell type annotations were assigned in a supervised manner by comparing the DEGs of each cluster together with the expression of established marker genes. Differential gene expression between DGUOK and control was performed using a pseudobulk approach. Raw counts were summed per cell type per subject, and differential expression was tested using edgeR's quasi-likelihood framework, with TMM normalization. A two-group model was fitted per cell type, with control as the reference, and results were aggregated across all cell types.

### Trajectory inference and gene expression dynamics analysis

Downstream trajectory analyses were carried out in Python v3.11.11 using Scanpy v1.11.5 and Palantir v1.4.3. For reconstruction of the aberrant basaloid trajectory[5], cells were reclustered with the Leiden algorithm (resolution = 0.7). Trajectory topology was then inferred with Partition-based Graph Abstraction (PAGA), using Leiden clusters as grouping variables. Edges with low confidence were removed by applying a connectivity threshold of 0.25, and the resulting abstracted graph was used to initialize a force-directed layout for visualization. To focus specifically on the alveolar epithelial differentiation axis, AT1, AT2, Aberrant Basaloid, iAT1, and iAT2 populations were subsetted for pseudotime inference. Trajectory modeling was performed in Palantir using graph-based diffusion without imputation. Aberrant Basaloid cells were designated as the early progenitor/root state, while AT1 and AT2 populations were set as terminal fates. Representative root and terminal cells were selected based on maximal within-cluster connectivity in the kNN graph. Palantir was run with these defined early and terminal states (knn = 20, 2500 waypoints), producing pseudotime values, differentiation entropy, and fate probabilities toward AT1 and AT2 endpoints. Cells were assigned to terminal fates using a probability threshold of >0.5, and fate trajectories were visualized through graph embeddings and Palantir branch plots. Final fate labels were further refined to ensure lineage consistency: cells annotated as AT1 or iAT1 were restricted to the AT1 branch, and cells annotated as AT2 or iAT2 were restricted to the AT2 branch. To support robust modelling of gene expression dynamics, each fate trajectory was downsampled to a maximum of 1000 cells per epithelial subtype. Pseudotime values were unity normalized and inverted so that Aberrant Basaloid cells mapped to early pseudotime, while terminal AT1 and AT2 states corresponded to late pseudotime. Gene expression dynamics along each trajectory were modelled using tradeSeq v1.20.0. For each fate branch, 2000 highly variable genes were selected, and generalized additive models were fit with negative binomial regression (fitGAM) using three knots. Association testing was then performed to identify genes with significant variation across pseudotime, with Benjamini–Hochberg correction applied to control the false discovery rate (adjusted $p < 0.05$).

**Spatial Validation**

The procedure has been described previously[6-8]. Slides were prepared as 2 µm thick tissue sections. Rehydration was done through a xylene (Carl Roth, Cat. CN80.1) and ethanol (Otto Fischar, Cat. 27690) series. Tissue decrosslinking was performed at 95°C in ddH2O with 1 % Antigen Unmasking Solution (Vector Laboratories, Cat. H3301) for 20 min. Slides were cooled down to RT for 20 min in 1X PBS (Chemsolute, Cat. 8418). Blocking of unspecific binding sides was reduced through incubation with 2.5% v/v normal horse serum (Vector Laboratories, Cat. S-2012) for 20 min. Incubation with primary antibodies was done for 1 h and tissue was rinsed twice with PBS for 2 min afterwards. DyLight™ 488 Anti-mouse-IgG and DyLight™ 594 Anti-rabbit-IgG secondary antibodies, both contained in the VectaFluor™ Duet Immunofluorescence Double Labeling Kit (Vector Laboratories, Cat. DK-8828), were applied to the slides for 1 h with a subsequent 1X PBS washing step as before. For three primary antibody panels, slides were incubated with pre-conjugated antibodies for 1 h, followed by a 1X PBS wash.

**Supplemental Table S2: Antibodies and respective dilutions used for multiple immunohistofluorescent antigen staining:**

| Target | Clonality (Clone) | Dilution | Manufacturer (Catatalog No.) |
|---|---|---|---|
| AGER | Monoclonal (A11) | 1:25 | Santa Cruz Biotechnology Inc. (sc-80652) |
| COX2 | Monoclonal (COX229) | 1:100 | Invitrogen (35-8200) |
| CTSE | Polyclonal | 1:100 | Merck/Sigma-Aldrich (HPA012940) |
| ITGAVB6 | Monoclonal EM05201) | 1:50 | Merck/Sigma-Aldrich (ZRB1104) |
| KRT17 | Monoclonal (E-4) | 1:25 | Santa Cruz Biotechnology Inc. (sc-393002 AF647) |
| LAMP3 | Polyclonal | 1:200 | Biotechne (AF4087) |
| SFTPC | Monoclonal (H-8) | 1:100 | Santa Cruz Biotechnology Inc. (sc-518029) |
| Mouse-IgG Rabbit-IgG | Polyclonal | Ready to use | VectaFluor Duet Immunofluorescence Double Labeling Kit: Vector Laboratories (DK-8828) |

The Vector TrueView Autofluorescence Quenching Kit (Vector Laboratories, Cat. SP-8400-15) was applied to each slide according to manufacturer's instructions to reduce background fluorescence, with 1X PBS rinsing afterward. DAPI-containing Antifade Mounting Medium (Vector Laboratories, Cat. H-1800-10) was used for coverslipping. Slides were stored in the dark at 4°C until imaging.

**RNAscope: RNA in situ hybridization**

The procedure has been described previously[6-8]. Slides were prepared as 2 μm thick tissue sections and incubated for 1 h at 60°C in an oven. Slides were rehydrated through 2x 5 min incubation in xylene and 2x 2 min incubation in 100% ethanol. Slides were dried in an oven for 5 min at 60°C. Afterwards, slides were treated with Hydrogen Peroxide (ACD, Cat. 322000) for 10 min at RT. Slides were rinsed in ddH2O by dipping the slides in and out ten times, twice. Slides were immersed in a pre-heated 1x target retrieval buffer (ACD, Cat. 322000) at 95°C for 30 min, twice rinsed in ddH2O for 1 min, and transferred to 100% ethanol for 1 min. Slides were removed from ethanol and ethanol was removed by drying for 5 min at 60°C. A hydrophobic barrier around the tissue was created using the intended pen (Vector Laboratory, Cat. H-4000). Tissue was digested using RNAscope Protease Plus (ACD, Cat. 322381) for 30 min at 40°C. Here and in between subsequent incubation periods, the slides were rinsed with 1x RNAscope Wash Buffer (ACD, Cat. 310091) for 2 min, twice. RNAscope probes (Bio-Techne, Cat. 401891, 401891-C2, 476341-C3) were mixed in the ratio 50:1:1 (C1:C2:C3) and pre-heated to 40°C for 10 min. The probe mix was applied to the slides for a 2 h at 40°C. For an optional overnight stopping point at RT, 5x SSC (Life Technologies, Cat. AM9770) was applied to the slides. The next day, incubations with RNAscope Multiplex FL v2 Amp 1, 2 and 3 were done in order at 40°C for 30 min, 30 min and 15 min, respectively. The Opal fluorophores (Akoya Biosciences, Cat. FP1487001KT, FP1488001KT, FP1495001KT, FP1497001KT) were reconstituted according to manufacturer's instructions and diluted 1:1000 in TSA Buffer (ACD, Cat. 322809). The fluorescence signal was developed for each channel independently through the following procedure: Incubation with RNAscope Multiplex FL v2 HRP-C1 (provided in ACD, Cat. 323110) for 15 min at 40°C, incubation with one specific Opal Fluorophore assigned to the channel for 30 min at 40°C, and incubation with RNAscope Multiplex FL v2 HRP-Blocker (provided in ACD, Cat. 323110) for 15 min at 40°C. Autofluorescence was quenched using the TrueBlack Lipofuscin Autofluorescence Quenching Solution (Biotium, Cat. B-23007) diluted 19:1 in 70% Ethanol for 1 min. Slides were stained with DAPI (ACD, part of Cat. 323110) for 15 s. Coverslipping was done with Prolong Gold Antifade Mounting Medium (Thermo Fisher Scientific, Cat. P36930). Slides were stored at 4°C in the dark until imaging.

#### Immunofluorescence Microscopy

Slides were imaged using the Zeiss Axio Scan 7 equipped with a 20x Plan-Apochromat 20x/0.8 27M objective. Emission was filtered through 96 HE BFP 450/40, 38 eGFP 525/50, 43 HE DsRed 605/70, 26 Alexa Fluor 660 685/50, and 50 Cy 5 690/50 broad pass filters.

#### Mitochondrial Respiration Analysis Using the Agilent Seahorse HS mini Analyzer

Mitochondrial respiration was assessed in fibroblasts of patient P1, P3 and P4 using the Agilent Seahorse HS mini Analyzer according to the manufacturer's instructions. Cells (20,000/well) were seeded in standard culture medium (see section cell culture) in Seahorse XFp microplates and incubated overnight. Sensor cartridges were hydrated overnight in sterile distilled water at 37 °C without $CO_2$. On the day of the assay, cells were washed and equilibrated for 1 hour in Seahorse XF assay medium (XF DMEM with 10 mM glucose, 1 mM pyruvate, and 2 mM glutamine, pH 7.4, Agilent Technologies, US) at 37°C in a non-$CO_2$ incubator. Sensor cartridges were transferred into calibrant solution and also incubated for 1 hour prior to the assay at 37°C without CO2. The Seahorse XFp Cell Mito Stress Test Kit (Agilent Technologies, US) was performed according to manufacturer's protocol by sequential injection of oligomycin (1.5 µM), FCCP (2 µM), and rotenone/antimycin A (0.5 µM each). Oxygen consumption rate (OCR) and extracellular acidification rate (ECAR) were measured in three cycles at baseline and after each subsequent injection. Data were analyzed using Seahorse Analytics software to calculate mitochondrial parameters like basal respiration, maximal respiration, proton leak, spare capacity, ATP production and non-mitochondrial respiration and normalized to cell number. Experiments were conducted in at least three independent replicates.

#### Lung biopsy and histology

Lung tissue biopsies have been sampled by video-assisted thoracoscopic surgery from patients P1, P2, P3 and P4, and by transbronchial cryobiopsy from patients P3 and P4. In addition, tissue obtained from the explanted lung of patient P1 was used. Samples were formalin fixed, paraffin embedded, and stained with hematoxylin-eosin, periodic acid–Schiff, and Masson's trichrome staining. In addition, samples were taken for transmission electron microscopy from severely affected areas (periphery of lung explant) and areas that appeared from a macroscopically point of view unaffected (more central region of the lung). Fixation and processing of these samples was performed in analogue to the cell pellets and is outlined in the next section.

#### Electron microscopy

Fibroblasts from patients P1, P3, and P4, as well as from two healthy relatives (the mother of P1 and the father of P4), were expanded in T75 flasks and harvested by trypsinization at 90% confluence. The cell pellets were fixed by a solution of 1.5% glutaraldehyde, 1.5% paraformaldehyde in 0.15 M HEPES buffer, treated with half-saturated uranylacetate solution, post-fixed with 4% OsO4 and dehydrated using a series of ascending acetone-concentrations from 70% to 100%. Finally, cell pellets were embedded in Epoxy resin. Ultrathin sections of 60 nm thickness were cut using an ultramicrotome (Leica, RM2265, Nussloch, Germany) and finally investigated by transmission electron microscopy (Morgagni, FEI, Eindhoven, The Netherland) equipped with a digital camera (Olympus Soft Imaging Solution, Münster, Germany).

### II. Supplementary Figures

#### Supplementary Figure S1. Analysis of Mitochondrial Respiration and Enzyme Activity

**A**

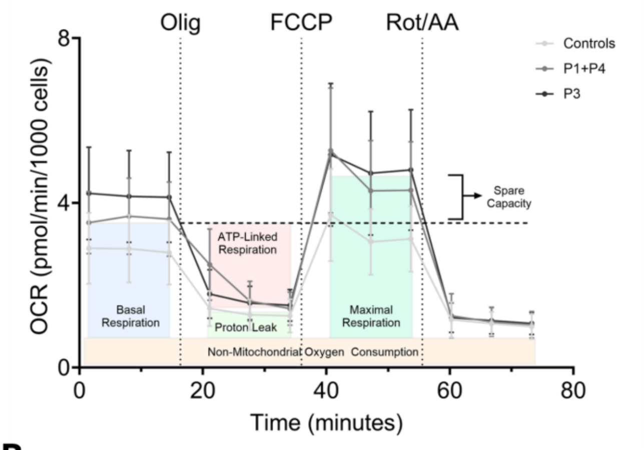

**B**

| | FB - P1 | FB - P3 | FB - P4 | FB – Ref. range | Muscle - P3 | Muscle - Ref. range |
|---|---|---|---|---|---|---|
| **Komplex I+III** | 92 | 79 | 56 | 61-157 | 26 | 18-88 |
| **Komplex II+III** | 7 | 4 | 2 | 4-13 | 3 | 5-83 |
| **Komplex IV** | 64 | 40 | 41 | 27-92 | 0 | 13-94 |
| **Komplex V** | 170 | 113 | 133 | 71-241 | 315 | 200-860 |
| **Citratsynthasis** | 77 | 82 | 69 | 50-155 | 79 | 61-114 |

Panel A: Oxygen consumption rate (OCR; pmol O2/min/1000 cells) in primary fibroblasts from controls (light grey), P1 combined with P4 (grey), and severely affected P3 (black) was determined by Cell Mito Stress test on a Seahorse XF HS mini extracellular flux analyzer. Three measurement cycles were taken: one for basal respiration (without any injection), as well as after each injection of Olig (Olygomycin), FCCP (Carbonylcyanide-4-(trifluormethoxy)phenylhydrazone) and Rot/AA (Rotenone / Antimycine A). After each assay cell number was determined and data was normalized to OCR/1000 cells. All data are presented as mean and SD from at least three independent experiments in triplicates (number of pooled wells: controls n=12, P1+P4 n=8+3, P3 n=9). Panel B: Respiratory chain complex activities in cultured skin fibroblasts (FB) from patients 1, 3 and 4 and respiratory chain complex activities in a muscle specimen from patient 3. Citrate synthase as a mitochondrial marker was in the reference range. Activities are in nmol/min per mg protein.

**Supplementary Figure S2. Transcriptional changes in epithelial cells**

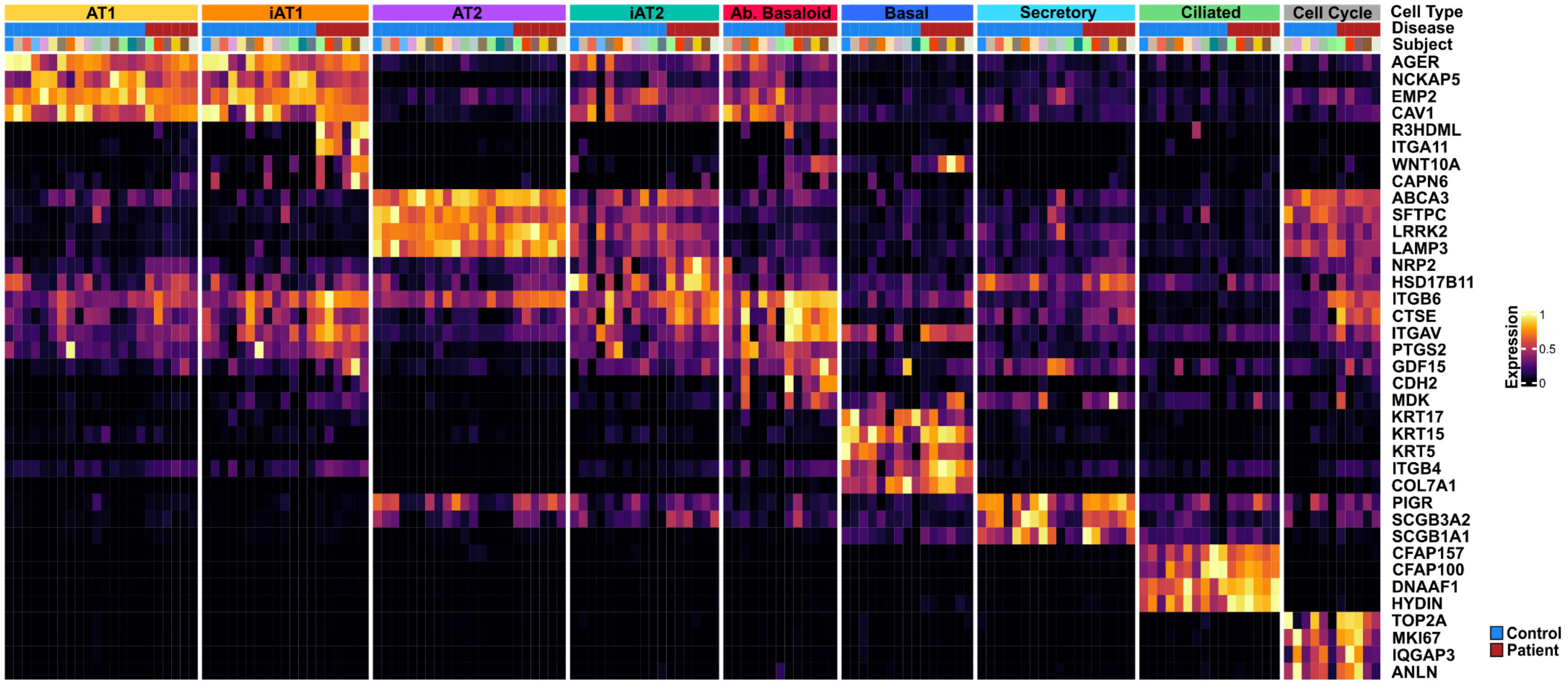

This heat map presents the normalized gene-expression values annotated by epithelial cell type and disease status.

**Supplementary Figure S3. Expansion of Profibrotic and Elastofibrotic Fibroblasts with associated transcriptional changes**

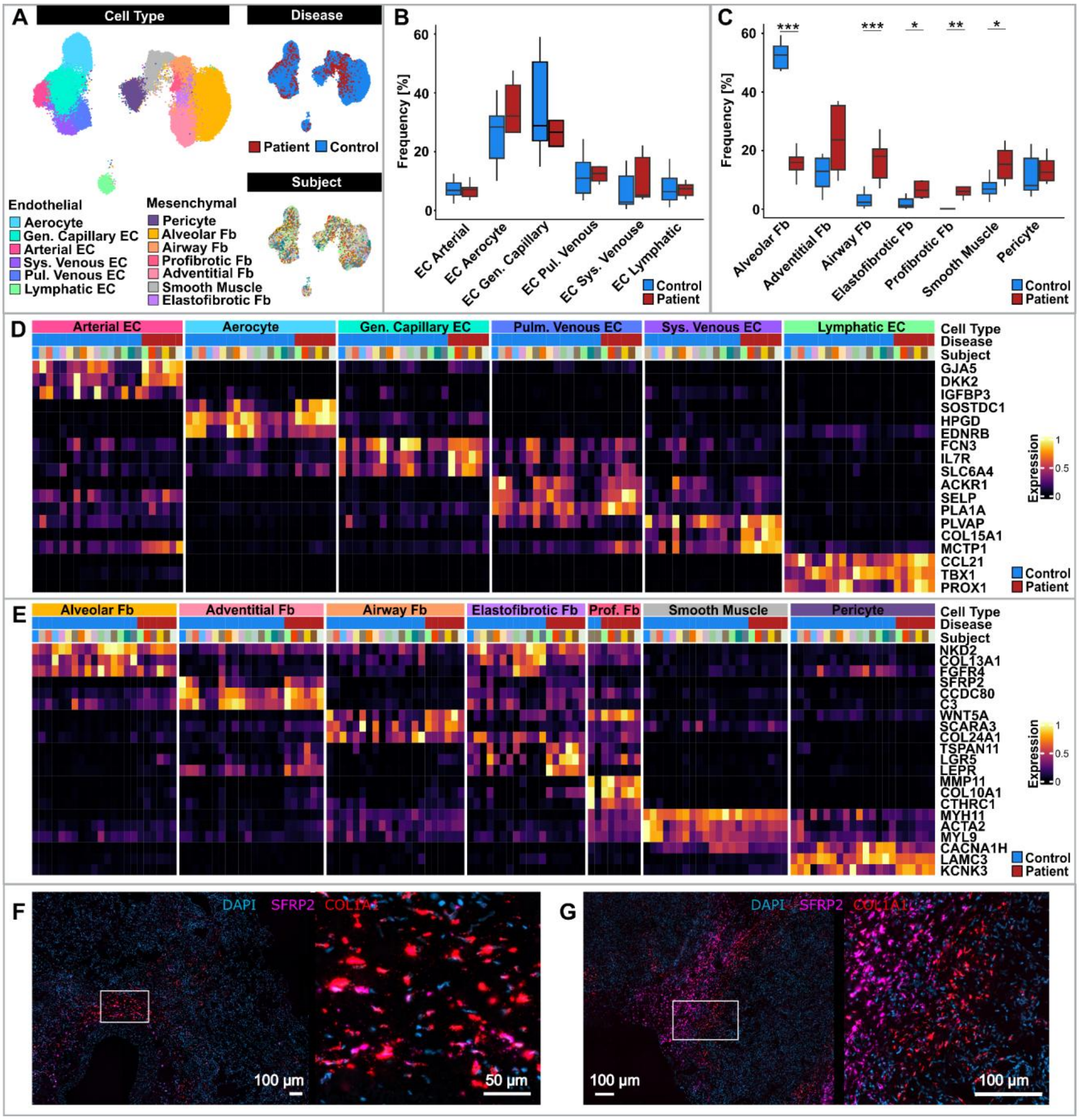

Panel A shows three UMAPs of endothelial and mesenchymal cells, annotated by cell type, disease, and subject. Panel B depicts the relative frequencies of endothelial cell types between PPFE in DGUOK-associated mitochondriopathy and controls, showing no significant differences. Box plots show the median, interquartile range, and full range of values Asterisks indicate statistical significance (*P<0.05; **P<0.01; ***P<0.001). Panel C illustrates the significant relative increase in Airway Fibroblasts (Fb), Elastofibrotic Fb, Profibrotic Fb, and Smooth Muscle cells, accompanied by a decrease in Alveolar Fb. Box plots show the median, interquartile range, and full range of values Asterisks indicate statistical significance (*P<0.05; **P<0.01; ***P<0.001). Panels D and E show heat maps of normalized gene-expression values, annotated by cell type and disease status. Panels F and G depict COL1A1-positive fibroblasts adjacent to Aberrant Basaloid cells, with SFRP2-positive fibroblasts, some of which co-express COL1A1.

**Supplementary Figure S4. Change in lung resident immune frequencies and gene expression**

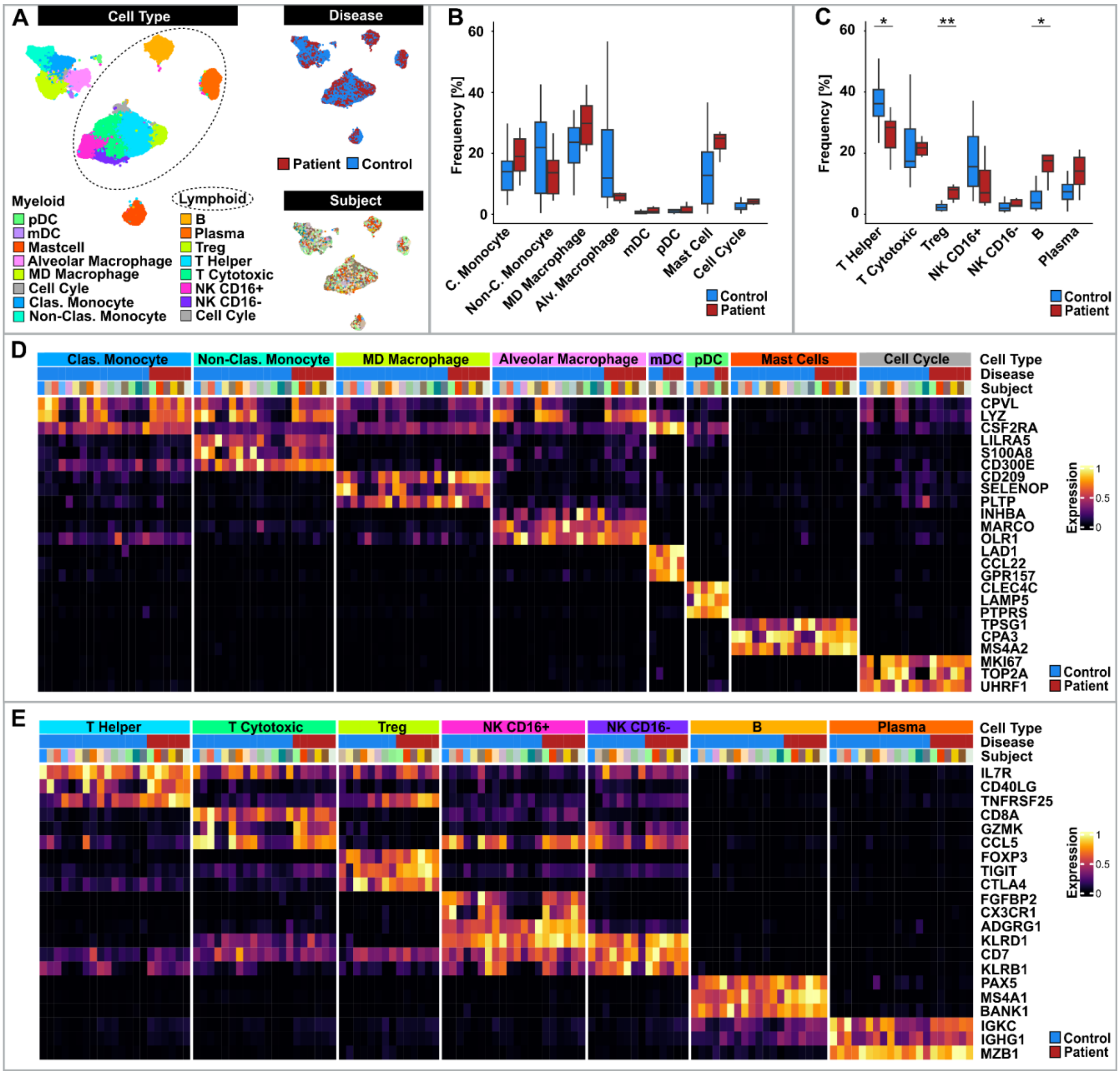

Panel A shows three UMAPs of myeloid and lymphoid cells, annotated by cell type, disease, and subject. Panel B depicts the relative frequencies of myeloid cell types between PPFE in DGUOK-associated mitochondriopathy and controls, showing no significant differences. Box plots show the median, interquartile range, and full range of values Asterisks indicate statistical significance (*P<0.05; **P<0.01). Panel C illustrates a significant relative increase in Tregs, and B cells with decrease in T Helper cells. Box plots show the median, interquartile range, and full range of values Asterisks indicate statistical significance (*P<0.05; **P<0.01; ***P<0.001). Panels D and E show heat maps of normalized gene-expression values, annotated by cell type and disease status in myeloid and lymphoid cell lineages, respectively.

**Supplementary Figure S5. Degenerated mitochondria in AT2 cells**

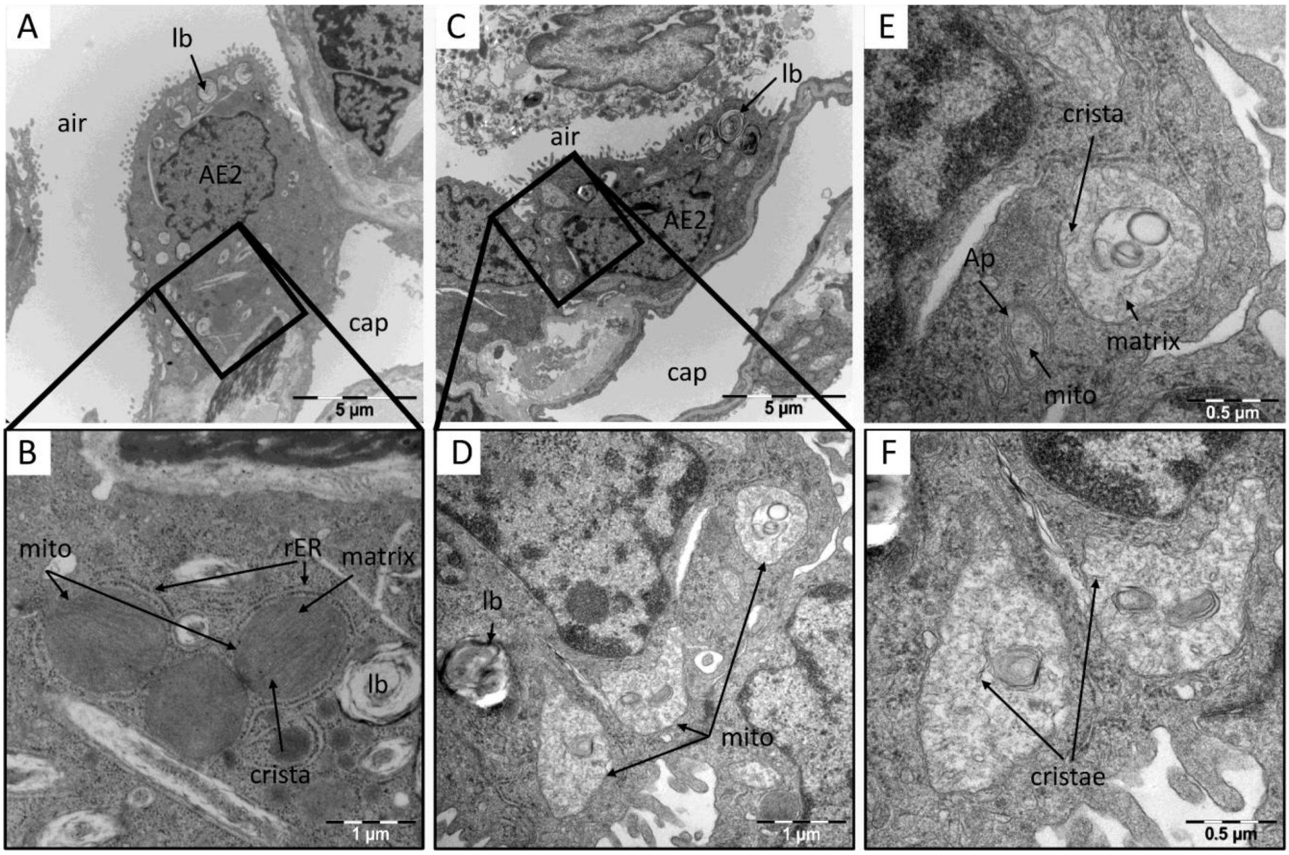

Panel A demonstrates an overview of an AT2 cell in a region with maintained acinar microarchitecture and absence of fibrosis of a lung explant diagnosed as Idiopathic Pulmonary Fibrosis. In Panel B details are illustrated showing enlarged mitochondria with maintained mitochondrial ultrastructure as characterized by two layers of biomembranes forming the inner and outer mitochondrial membrane, slim and densely packed cristae and in the space between the cristae electron dense mitochondrial matrix. Near to the mitochondria rough endoplasmic reticulum (rER) and a lamellar body (lb) can be found. Panel C illustrates a representative AT2 cell in a region of the lung of maintained acinar microarchitecture and absence of fibrosis in a lung explant suffering from DGUOK-deficiency. Panels D-F show details of the ultrastructure of the AT2 cell with a focus on the mitochondria (mito). In general, these are enlarged with diameters of 1 µm and more. While the enveloping double layer of biomembranes is maintained, there is a rarefaction of normal appearing slim cristae which can be identified as remnants at best. Other cristae appear to be dilatated, fragmented and thus subject of cristolysis. Moreover, loss of the mitochondrial matrix can be seen in many mitochondria. Although the process of mitophagy is a very rare event in AT2 cells an autophagosome (Ap) containing a fragment of an mitochondrium (mito) is demonstrated in Panel E.

**Supplementary Figure S6. Ultrastructure of cultured fibroblasts do not show differences between *DGUOK* deficiency and healthy controls**

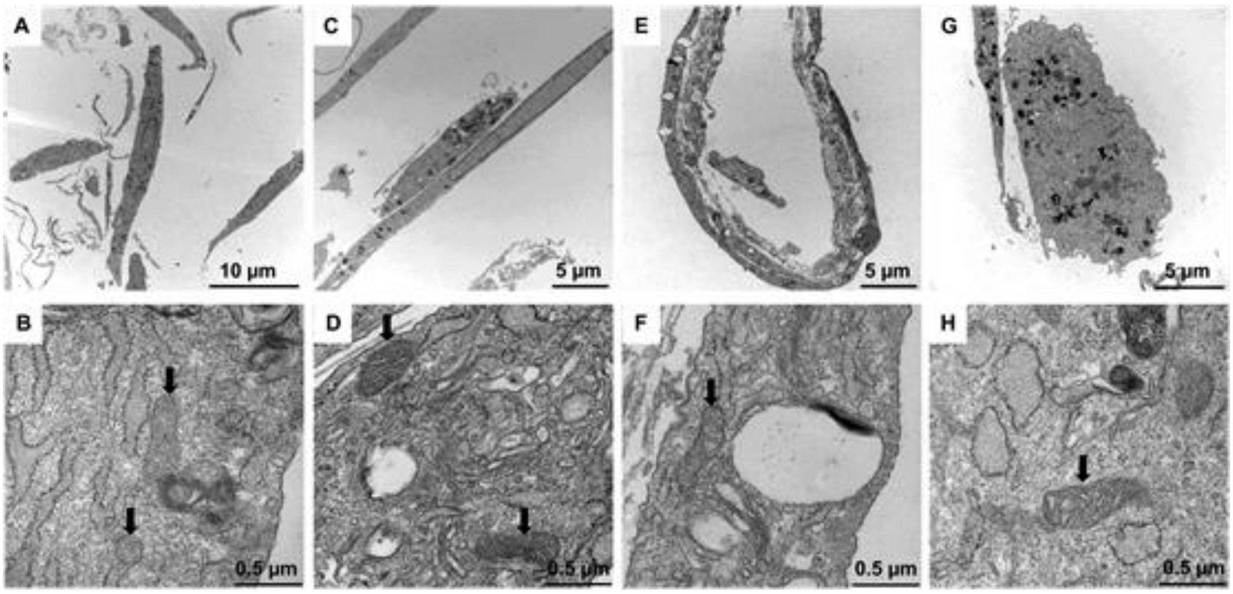

The upper row demonstrates overview images of the ultrastructure of cultured fibroblast and the lower row details at higher magnification with a focus on mitochondria (arrows). Panels A-F show fibroblasts of patients suffering from *DGUOK*-deficiency while Panel G and H are from a healthy control. Immediately after removal from the incubator, cultures of fibroblast were fixed. The culture medium was replaced by a solution of 1.5% glutaraldehyde, 1.5% paraformaldehyde in 0.15 M HEPES buffer. In general, the sizes of the mitochondria were in all patients in a normal range. No signs of mitochondrial degeneration were observed. The cristae were slim and regularly arranged. The mitochondrial matrix was electron dense in all cases.

### III. Supplementary Tables

#### Supplementary Table S3. Clinical characteristics

| | Patient 1 | Patient 2 | Patient 3 | Patient 4 | Patient 5 | Patient 6 | Patient 7 | Patient 8 | Patient 9 |
|---|---|---|---|---|---|---|---|---|---|
| Sex | Male | Female | Male | Female | Female | Male | Male | Male | Female |
| Parental consanguinity | yes | Yes | Yes | No | Yes | Yes | Yes | Yes | Yes |
| Primary manifestation | Insufficient weight gain and exertional dyspnea | Insufficient weight gain, exertional dyspnea | Liver failure | Insufficient weight gain and exertional dyspnea | Liver failure | Exertional dyspnea and muscle complaints | Exertional dyspnea | Exertional dyspnea and muscle complaints | Exertional dyspnea |
| Age at onset [years] | 5 - 15 years | 5 - 15 years | <5 years | 5 - 15 years | < 5 years | 18 - 30 years | 18 - 30 years | 18 - 30 years | 18 - 30 years |
| Age at first evaluation | 10 – 17 years | 10 – 17 years | <10 years | 10 – 17 years | < 10 years | > 30 years | > 30 years | 18 - 30 years | 18 - 30 years |
| Detected DGUOK variants (ENST00000264093.9) | c.592-26A>G/ c.592-26A>G | c.592-26A>G/ c.592-26A>G | c.592-26A>G/ c.313C>T | c.592-26A>G/ c.592-26A>G | c.763_766dup/ c.763_766dup | c.592-26A>G/ c.592-26A>G | c.592-26A>G/ c.592-26A>G | c.592-26A>G/ c.592-26A>G | c.592-26A>G/ c.592-26A>G |
| **Liver involvement** | | | | | | | | | |
| Cholestasis | No | No | Yes | No | Yes | No | No | No | No |
| Jaundice | No | No | Yes | No | Yes | No | No | No | No |
| Coagulopathy | No | No | Yes | No | Yes | No | No | No | No |
| Hepatomegaly | No | yes | No | No | Yes | No | No | No | No |
| Ascites | No | No | Yes | No | Yes | No | No | No | No |
| Hematemesis | No | No | No | No | No | No | No | No | No |
| Portal hypertension | No | No | Yes | No | Yes | No | No | No | No |
| Hepatocellular carcinoma | No | No | No | No | No | No | No | No | No |
| Liver transplantation | No | No | Yes | No | Yes | No | No | No | No |
| **Renal involvement** | | | | | | | | | |
| Proteinuria | Yes | Yes | Yes | No | NA | Yes | No | No | No |
| Renal insufficiency | No | No | No | No | No | Yes | No | No | No |
| **Skeletal muscle involvement** | | | | | | | | | |

| | | | | | | | | | |
|---|---|---|---|---|---|---|---|---|---|
| Muscle weakness | Yes | No | Yes | Yes | NA | Yes | Yes | Yes | No |
| Truncal weakness | Yes | No | Yes | Yes | NA | No | No | No | No |
| Muscle pain | No | No | No | No | NA | Yes | Yes | Yes | No |
| Muscle atrophy | No | No | No | No | NA | Both hands | Yes | Yes | No |
| **Respiratory involvement** | | | | | | | | | |
| Respiratory distress / dyspnea | Yes | Yes | Yes | Yes | Yes | Yes | Yes | Yes | No |
| History of pneumothorax | Yes, chronic | No (only iatrogenic after biopsy) | persistent after biopsy | Yes, chronic | No | No | Yes | Yes | No |
| Chronic cough | Yes | No | Yes | Yes | No | Yes | Yes | No | No |
| Initial FVC in L (% of predicted) | 0.9 (19) | 1.16 (39) | 0.34 (12) | 0.55L (16) | NA | 1.18 (25) | NA | NA | NA |
| FEV1 in L (% of predicted) | 0.77 (19) | 1.16 (43) | NA | NA | NA | 1.10 (28) | NA | NA | NA |
| DLCO in % of predicted | NA | 53% | NA | NA | NA | 34 | NA | NA | NA |
| **Cardiovascular involvement** | | | | | | | | | |
| Cardiomyopathy | No | No | No | No | No | No | No | No | No |
| Pulmonary hypertension | No | No | No | No | Yes | No | No | No | No |
| Arterial Hypertension | No | No | No | No | No | Yes | No | No | No |
| **Gastrointestinal involvement** | | | | | | | | | |
| Vomiting | No | No | Yes | No | Yes | No | No | No | No |
| **Weight or growth abnormalities** | | | | | | | | | |
| Height [cm] | 170 | 150 | 87 | 158.5 | 66 | 173 | 170 | 178 | 165 |
| Weight [kg] | 40 | 37 | 10 | 30.8 | 5 | 61 | 40 | 55 | 42 |
| Age at height & weight measurement | 17 | 15 | 2.75 | 15 | 0.8 | 36 | 33 | 27 | 22 |
| BMI | 13.8 | 16.4 | 13.2 | 12.3 | 11.4 | 20.4 | 13.8 | 17.4 | 15.4 |
| **Ocular involvement** | | | | | | | | | |
| Ophthalmoplegia | No | No | No | No | No | No | No | No | No |
| Ptosis | No | No | No | No | No | No | No | No | No |
| Visual impairment | No | No | Yes (Refractive error) | No | No | No | No | Yes (Refractiv e error) | No |
| **Neurological involvement** | | | | | | | | | |
| Nystagmus | No | No | No | No | No | No | No | No | No |
| Global developmental delay | No | No | Yes | No | Yes | No | No | No | No |

| | | | | | | | | | |
|---|---|---|---|---|---|---|---|---|---|
| Seizures/abnormal EEG | No | No | No | No | No | No | No | No | No |
| Hyporeflexia | NA | Yes | Yes | NA | NA | Yes | NA | NA | NA |
| Encephalopathy | No | No | No | No | Yes in liver failure | No | No | No | No |
| Peripheral neuropathy | Yes | Yes | Yes | Yes | No | Yes | No | No | No |
| Strabismus | No | No | No | No | NA | No | No | No | No |
| Dysphagia | No | No | Yes | No | NA | No | Yes | No | No |
| **Other symptoms** | | | | | | | | | |
| Suprasternal notch | yes | yes | yes | Yes | NA | Yes | Yes | Yes | Yes |
| Fatigue | yes | Yes | Yes | Yes | NA | Yes | Yes | No | No |
| Other | | Mild hypothyroid-dism | Osteopo-rosis | | Pseudohypo-aldostero-nism, therapy-resistant dermatitis | | Chronic venous insufficienc y | Frequent headache s, Gyneco-mastia | |
| **Laboratory findings** | | | | | | | | | |
| Elevated Lactate | Yes | No | Yes (in liver failure) | Yes | Yes | No | NA | NA | NA |
| Low blood glucose | No | No | Yes | No | Yes | No | No | No | No |
| Elevated ALT | No | No | No | Yes | Yes | No | Yes | Yes | No |
| Elevated AST | No | Yes | Yes | Yes | Yes | No | Yes | Yes | No |
| Elevated AFP | NA | NA | Yes | NA | NA | NA | NA | NA | NA |
| Elevated GGT | Yes | Yes | Yes | Yes | Yes | No | No | No | NA |
| Elevated tyrosine | NA | No | No | NA | Yes | NA | NA | NA | NA |
| Elevated ammonia | NA | NA | Yes | NA | Yes | NA | NA | NA | NA |
| Elevated ferritin | No | No | No | No | Yes | No | NA | NA | No |
| Low albumin | No | No | Yes | No | Yes | No | No | No | NA |
| Elevated alanine | NA | No | No | NA | Yes | NA | NA | NA | NA |
| Elevated CK | No | Yes | No | Yes | Yes | No | Yes | Yes | Yes |
| Elevated AP | NA | No | No | No | Yes | No | No | No | NA |
| Elevated methionine | NA | No | No | No | Yes | NA | NA | NA | NA |
| Elevated NT-proBNP | No | No | No | No | Yes | No | NA | NA | NA |
| **Status** | | | | | | | | | |
| Current status | Alive | Alive | Deceased | Alive | Deceased | Alive | Alive | Alive | Alive |

| | | | | | | | | | |
|---|---|---|---|---|---|---|---|---|---|
| If deceased, age at death [years] | NA | NA | 9 | NA | 5 | NA | NA | NA | NA |
